# Time-of-Day Variation in Subjective Mood Among Lumosity Users

**DOI:** 10.1101/2022.05.12.22275014

**Authors:** Raymond A. Mendoza, Fabian-Xosé Fernandez, Andrew S. Tubbs, Michael L. Perlis, Michael A. Grandner

**Author notes:** Co-corresponding authors Please address correspondence to either: Michael L. Perlis, PhD, Behavioral Sleep Medicine Program, Department of Psychiatry, University of Pennsylvania, PA, USA, Michael A. Grandner, PhD, Sleep and Health Research Program, Department of Psychiatry, University of Arizona, AZ, USA. Co-first authors (Raymond A. Mendoza, Fabian-Xosé Fernandez).

## Abstract

Human mood varies according to both endogenous and situational factors. Analyzing patterns of online media engagement can provide additional insights into a third dimension of mood potentially involving time-of-day factors. Using data collected through the online brain-training app, Lumosity, we report real-world trends in mood expression among 500,000 respondents. Endorsements of positive mood by Lumosity gamers peaked midday, while endorsements of negative mood peaked overnight. These data suggest that there may be inherent bias towards negative mood when individuals are awake at night. More broadly, they suggest that internet data-trails have the potential to uncover fundamental aspects of human behavior.

## 1. Introduction

Mood varies in its expression across the day and night (Bolvin et al., 1997; Dzogang et al., 2017; Emens et al., 2020; Golder and Macy, 2011; Monk et al., 1992). Mood states are distinguished by their display of positive versus negative affect (Norman et al., 2011; Norris et al., 2010; Watson et al., 1999). Positive affect reflects a positive internal disposition whereas negative affect reflects the opposite. Both affects have been shown to follow their own circadian trajectories, which appear to travel independently of one another (Adan and Sánchez-Turet, 2001; Clark et al., 1989; Hasler et al., 2008; Miller et al., 2015; Murray et al., 2009; Porto et al., 2006; Stone et al., 2006; Vittengl and Holt, 1998). As such, a change in positive affect does not portend changes in negative affect (or vice versa).

Previous in-lab studies have documented diurnal rhythms of mood that are under the control of the endogenous circadian pacemaker (Bolvin et al., 1997; Emens et al., 2020; Monk et al., 1992). More recently, diurnal mood variation has been studied via online data exploration (mostly on Twitter) by coding linguistic expressions into their respective affective states. Once compiled, these data contain a much greater sample size than those collected under experimental settings owing to (1) the accessibility and quantity of internet data versus (2) the relative difficulties of conducting multi-day studies in isolation facilities. Interestingly, under both research designs, subjective indices of positive affect tend to rise and peak during the middle of the day and decline thereafter throughout the night. Conversely, subjective indices of negative affect tend to rise and peak in the dead of night and then diminish starting at daybreak (Dzogang et al., 2017; Dzogang et al., 2018; Golder and Macy, 2011).

Studies analyzing data from online media platforms are subjective in the sense that they are reported through the language of the users. This “indirect” expression (e.g., linguistic coding of social media posts) will reflect mood to varying degrees based on factors like culture. By contrast, collection of survey data directly inquiring about mood can better confirm realworld circadian trends. Lumosity is a brain-training application available on mobile devices with the goal of improving cognitive skills in everyday life. The application has a supplemental mood assessment feature, which is prompted upon logging in and asks the user to rate their mood from a scale at the time of answering. Here, we report the first use of these data to examine time trends in mood expression. Based on previous in-lab work and online data exploration, it was hypothesized that the peak expression of positive affect among Lumosity users would occur in the middle of the day, while peak expression of negative affect would occur in the middle of the night.

## 2. Methods

Deidentified summary data from N=493,758 unique Lumosity users were made available by Lumos Labs, Inc. (San Francisco, CA) at our request. As part of the app’s basic functionality, all participants rated their mood when they logged in. Hour of assessment was recorded. Mood was rated along a 5-point analog scale. Mean scores, as well as percent positive (4-5) or negative (0-1) were evaluated by hour. To determine whether an overall pattern exists, ANOVA was performed on mean score and chi-square tests were performed on percentage scores. Time of day was then binned in 4-hour increments (0:00-3:59, 4:00-7:59, 8:00-11:59, 12:00-15:59, 16:00-19:59, and 20:00-23:59) and similarly evaluated. A subsample of users was analyzed with 400 participants in each age (per) sex (per) hour bin (for a total of 76,800 unique users). The current study was determined to be exempt from human-subjects review by the University of Arizona Institutional Review Board.

## 3. Results

Mean mood ratings varied across the day (F=42.98, *p* < 0.0001) among Lumosity users, as did the proportion of both positive and negative ratings separately (*p* < 0.0001). Endorsements of positive mood peaked at approximately 15.00 (50.5%) and then slowly declined until 09.00 the next day (43.3%), at which point they began rising again (**Figure 1**, top panel). Endorsements of negative mood followed a reciprocal pattern, with a trough at 14.00 (7.9%) that rose slowly until 09.00 the next day (11.5%). When the 24-h period was divided into 6 equal bins, there was a significant difference noted among them (*p* < 0.0001), with all pairwise comparisons significant (*p* < 0.0001) except for 12:00-15:00 and 16:00-19:00. The presence of 24-h diurnal rhythms in both positive and negative mood endorsements was confirmed by a significant fit of each dataset to a sinusoidal curve with a period of 24 h (R^2^ values = 0.69-0.74, *ps* < 0.0001). None of these results changed in the subsample balanced for age and sex (data not shown). Proportions relative to maximum reports are displayed in the bottom panel of **Figure 1**.

**Figure 1.**
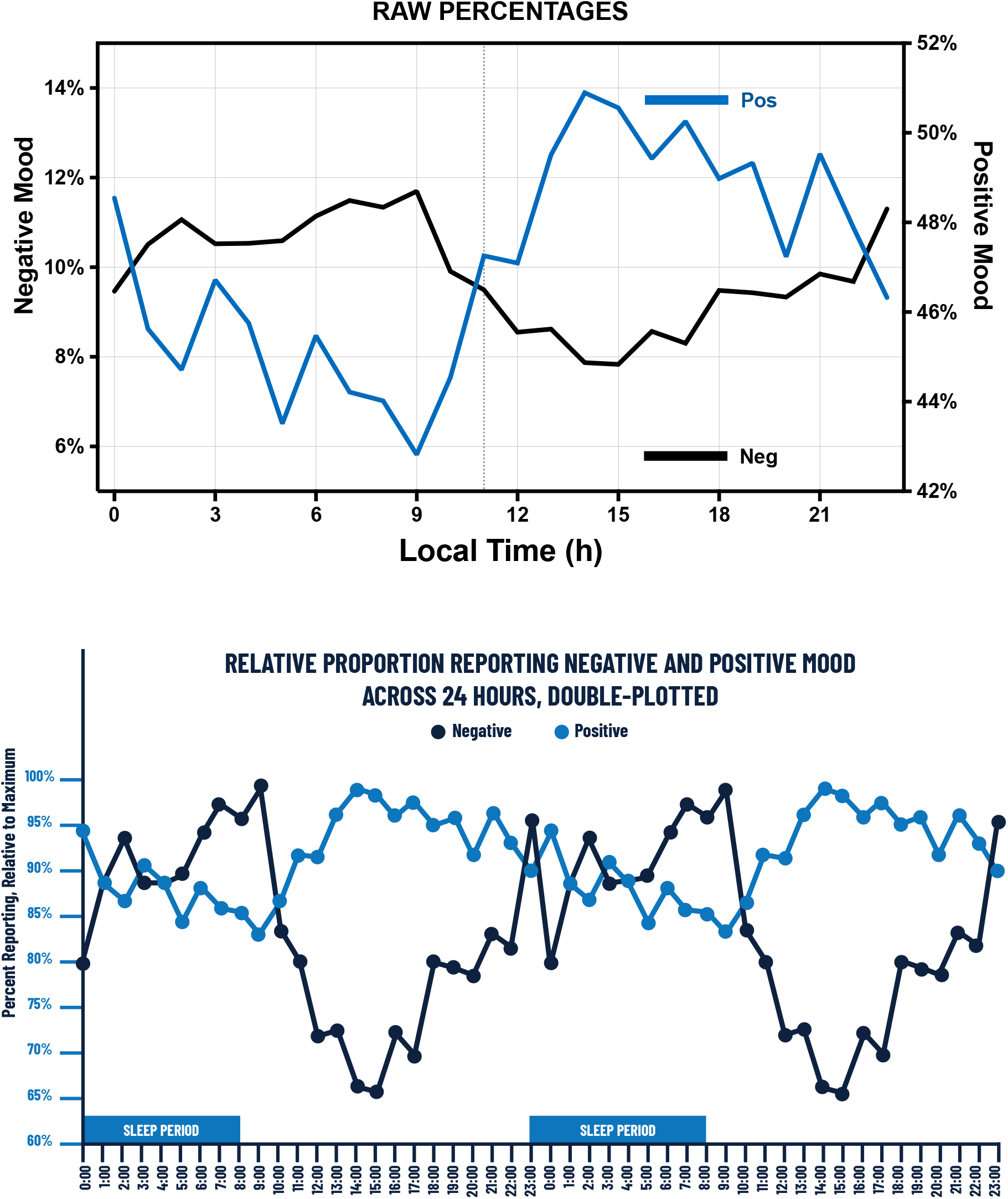
Percentage of positive and negative mood ratings among Lumosity users (top panel) relative to the maximum reporting period (bottom panel). Data are double-plotted to better visualize daily trends.

## 4. Discussion

Diurnal variation in reports of subjective mood were evident among Lumosity users, with positive affect peaking midday and negative affect peaking overnight. The trough of positive affect lingered throughout the night coinciding with the sleep period (with ratings rebounding in the morning), while the trough of negative affect was centered in the afternoon. These patterns—and their approximate timing—bear strong similarities to those documented during in-lab circadian and sleep deprivation experiments (e.g., Emens et al., 2020). The overnight peak in negative affect is of clinical interest because it may be one of several factors contributing to the higher risk of suicide during the circadian night (Tubbs et al., 2020b; Tubbs et al., 2021a; Tubbs et al., 2021b). It is possible that people who are awake in the middle of the night experience the full brunt of the peak in negative affect at a time when most are shielded from it because they are sleeping (Emens et al., 2020). Furthermore, sleep may serve a restorative function by which negative affect is “refreshed”; if so, nocturnal wakefulness would impede this recovery (Golder and Macy, 2011). Consistent with these general trains of thought, online data analysis has identified variations throughout the day in the timing of suicide-related posts on forums such as Reddit’s r/SuicideWatch. Post volume peaks in the early morning between 2:00–5:00 h (Dutta et al., 2021), at a time commensurate with the interval we have identified for when Lumosity users report maximal negative affect and minimal positive affect.

Diurnal rhythms of mood in the positive and negative dimensions have now been identified using multiple online data analysis strategies, including self-report and online linguistic coding. Twitter has been subject to most online study given its visible role in social media as well as its potential to accurately measure people’s personality in a naturalistic setting (Qiu et al., 2012). Various linguistic coding tools have been employed for analysis of Twitter posts, most prominently the LIWC (Linguistic Inquiry and Word Count). That said, other approaches in and outside of Twitter have been tested as well, producing complementary and in some cases alternative results. In one instance, online data exploration identified diurnal variations in musical preference on the streaming app, Spotify, where users showed significant preferences for low-valence (more ‘sad, depressed’) tracks during the night and early morning hours between 11pm and 6am (Heggli et al., 2021). These time trends match the general time trends uncovered for mood among Lumosity and Twitter users, and for suicide posts to Reddit. However, in another instance, emoji expression was shown to follow a pattern distinct from the one found for text-type patterns of mood (Mayor and Bietti, 2021), suggesting that there might be something inherently different about image expression of mood versus its text expression or its manifestation in music preference.

Various other factors are important to consider when discussing circadian rhythms of mood, such as sex, age, and chronotype. In the present study, significance remained across four-hour increments when controlling for age and sex, though we were unable to assess the contribution of chronotype. Still, the present findings add materially to the literature documenting circadian trends in mood expression across different analytical approaches and research designs. Future studies should continue to evaluate whether these differences manifest across continents and cultures, and to build more comprehensive linguistic lexicons by which to quantify the expression of positive and negative affect. Such efforts might contribute to the development of more effective overnight suicide intervention strategies (Kaladchibachi and Fernandez, 2018; Tubbs et al., 2020a).

## Data Availability

All data are property of Lumos Labs and are available upon reasonable request from the company.

## Acknowledgements

The authors were supported as follows: AST: DOD W81XWH-16-2-0003 (F-XF, MG) and NIH K24AG055602 (MLP). F-XF: Velux Stiftung (Project No. 1360). MLP: NIH K24AG055602 and R01AG054521. MAG: R01DA051321 and R01MD011600.

